# Oxidative stress and antioxidant defenses in mild cognitive impairment: a systematic review and meta-analysis

**DOI:** 10.1101/2021.11.22.21266698

**Authors:** Gallayaporn Nantachai, Asara Vasupanrajit, Chavit Tunvirachaisakul, Marco Solmi, Michael Maes

**Affiliations:** Department of Psychiatry, Faculty of Medicine, Chulalongkorn University, Bangkok, Thailand; Somdet Phra Sungharaj Nyanasumvara Geriatric Hospital, Department of Medical Services, Ministry of Public health, Chon Buri Province, Thailand; Cognitive Impairment and Dementia Research Unit, Department of Psychiatry, Faculty of Medicine, Chulalongkorn University, Bangkok, Thailand; Department of Psychiatry, University of Ottawa, Ontario, Canada; Department of Mental Health, The Ottawa Hospital, Ontario, Canada; Ottawa Hospital Research Institute (OHRI) Clinical Epidemiology Program University of Ottawa, Ottawa, Ontario; Early Psychosis: Interventions and Clinical-detection (EPIC) Lab, Institute of Psychiatry, Psychology & Neuroscience, Department of Psychosis Studies, King’s College London, London, United Kingdom; Centre for Innovation in Mental Health-Developmental Lab, School of Psychology, University of Southampton, and NHS Trust, Southampton, United Kingdom; IMPACT Strategic Research Center, Deakin University, Geelong, Australia; Department of Psychiatry, Medical University of Plovdiv, Plovdiv, Bulgaria

**Keywords:** Oxidative and nitrosative stress, antioxidants, biomarkers, neuro-immune, neurocognition

## Abstract

This study aims to systematically review and meta-analyze the nitro-oxidative stress (O&NS)/antioxidant (ANTIOX) ratio in the peripheral blood of people with mild cognitive impairment (MCI). We searched PubMed, Scopus, Google Scholar, and Web of Science for articles published from inception until July 31, 2021. Forty-six studies on 3.798 MCI individuals and 6.063 healthy controls were included. The O&NS/ANTIOX ratio was significantly higher in MCI than in controls with a Standardized Mean Difference (SMD)=0.378 (95% CI: 0.250; 0.506). MCI individuals showed increased lipid peroxidation (SMD=0.774, 95%CI: 4.416; 1.132) and O&NS-associated toxicity (SMD=0.621, CI: 0.377; 0.865) and reduced glutathione (GSH) defenses (SMD=0.725, 95%CI: 0.269; 1.182) as compared with controls. MCI was also accompanied by significantly increased homocysteine (SMD=0.320, CI: 0.059; 0.581), but not protein oxidation, and lowered non-vitamin (SMD=0.347, CI: 0.168; 0.527) and vitamin (SMD=0.564, CI: 0.129; 0.999) antioxidant defenses. The results show that MCI is at least in part due to increased neuro-oxidative toxicity and suggest that treatments targeting lipid peroxidation and the GSH system may be used to treat or prevent MCI.

## Introduction

Cognitive impairment often occurs with a prevalence of 10%-15% in elderly individuals aged over 65 years (Anderson, 2019). Amnestic Mild Cognitive Impairment (aMCI) is defined by milder deficits in neurocognitive functions, most notably episodic memory, language difficulties, poor learning ability, and problem solving, but without functional decline in basic activities of daily living (ADL) (Hemrungrojn et al., 2021; Petersen et al., 2010). The conversion rate from aMCI to Alzheimer’s disease is approximately 16.5 percent per year, but some patients (8%) with aMCI may revert to a normal state, indicating that some, but not all, aMCI patients are at increased risk of developing dementia (Petersen et al., 2010). While increasing age is the primary risk factor for aMCI, male sex, metabolic syndrome and “being less engaged in mental activities” may also play a role, while increased education, cognitive training, physical activity, and a Mediterranean diet are protective factors (Etgen et al., 2011; Hussin et al., 2019; Roberts and Knopman, 2013).

According to the oxidative stress theory of aging, aging-related functional losses are thought to be caused at least in part by a buildup of damage due to reactive oxygen and nitrogen species (RONS) (Liguori et al., 2018; Luo et al., 2020; Tan et al., 2018). Likewise, increased oxidative and nitrosative stress (O&NS) is linked to many age-related disorders, which are risk factors of aMCI, including hypertension and type-2 diabetes mellitus (T2DM) (Liguori et al., 2018; Maes and Tangwongchai, 2021). There is also evidence that age-associated damage due to increases in RONS‒induced O&NS damage to mitochondria are associated with neurodegenerative disease including Alzheimer’s disease (E Abdel Moneim, 2015; Islam, 2017).

A previous meta-analysis showed increased O&NS biomarkers, such as signs of increased lipid peroxidation and copper metabolism, in the peripheral blood of people with aMCI as compared with healthy controls (Schrag et al., 2013). Importantly, lipid peroxidation biomarkers may be used to detect MCI and the early stages of Alzheimer’s disease with adequate specificity and sensitivity (Arslan et al., 2020). Not only signs of lipid peroxidation but also increased protein oxidation as assessed with advanced oxidation protein products (AOPP), and plasma homocysteine (Hcy) levels may be increased in patients with MCI and Alzheimer’s disease (Ansari, 2016; Religa et al., 2003; Vergallo et al., 2018). Assays of O&NS biomarkers including isoprostanes may serve as biomarkers of MCI and potential early Alzheimer disease (García-Blanco et al., 2017).

Several antioxidant systems including the glutathione (GSH) system, superoxide dismutase (SOD), catalase (CAT), high density lipoprotein cholesterol (HDL-c), vitamins A, E and C, and carotenes, may protect against O&NS damage to lipids, proteins, DNA, and mitochondria (Maes et al., 2011) are reduced in aMCI (Schrag et al., 2013) and Alzheimer’s disease (da Silva et al., 2014). A first Meta-Analysis (García-Blanco et al., 2017; Schrag et al., 2013) detected reduced non-enzymatic antioxidant levels (including vitamins A, E and C, and carotenes) in aMCI patients, while another meta-analysis showed that α-tocopherol levels were lower in people with MCI versus healthy controls (Ashley et al., 2019). In aMCI and Alzheimer’s disease lowered folic acid levels are observed (Ma et al., 2017; Religa et al., 2003), while another study found that thiol (- SH) groups and iron reducing ability of plasma (FRAP) are lower in MCI than in controls (Vergallo et al., 2018). Maes and Tangwongchai (2021) reported that a composite score based on the Apolipoprotein E4 (ApoE4) genotype, reduced antioxidant levels (namely folic acid and albumin), and other biomarkers, hypertension and T2DM significantly discriminated aMCI individuals from normal controls and patients with Alzheimer’s disease. da Silva et al. (2014) showed that folate and vitamin A, B12, C, and E levels were significantly reduced in patients with Alzheimer’s disease as compared with controls.

Nevertheless, it has remained elusive a) whether the overall O&NS/antioxidant (O&NS/ANTIOX) ratio is increased in MCI, and b) which of the O&NS/ANTIOX profiles (e.g., lipid peroxidation, protein oxidation, homocysteine metabolism, the GSH system, iron status, vitamin, and non-vitamin antioxidants) contribute most to the O&NS pathophysiology of MCI. Hence, the current study was performed to systematically review and meta-analyze the O&NS and ANTIOX data in MCI *versus* healthy controls and to delineate the most important O&NS/ANTIOX profiles of MCI.

## Materials and Methods

The methodological approach was established in accordance with the Preferred Reporting Items for Systematic Reviews and Meta-Analyses (PRISMA) 2020 (Page et al., 2021), the Meta-Analyses of Observational Studies in Epidemiology (MOOSE) (Stroup et al., 2000), and the Cochrane Handbook for Systematic Reviews and Interventions (Higgins et al., 2019). We followed an a priori protocol that can be obtained from the last author. This study aimed to delineate O&NS and ANTIOX profiles of people with MCI *versus* normal controls and, therefore, we have grouped different biomarkers according to their established functions into synthesized scores which reflect O&NS and ANTIOX profiles. The different profiles are shown and described in **Electronic Supplementary File (ESF) Table 1**.

**Table 1.** Number of cases with Mild Cognitive Impairment (MCI) and healthy controls in the different meta-analyses, and side of 95% confidence intervals and standardized mean difference (SMD) with respect to zero SMD.

| Outcome profiles | n studies | Side of 95% confidence intervals |  |  |  | MCI participants | Control participants | Total number of participants |
| --- | --- | --- | --- | --- | --- | --- | --- | --- |
|  |  | < 0 | Overlap 0 and SMD < 0 | Overlap 0 and SMD > 0 | > 0 |  |  |  |
| O&NS/ANTIOX | 46 | 0 | 6 | 22 | 18 | 3.798 | 6.063 | 9.861 |
| O&NS | 36 | 1 | 3 | 15 | 13 | 2.325 | 4.257 | 6.582 |
| O&NS toxicity | 30 | 1 | 4 | 13 | 12 | 1.795 | 2.275 | 4.070 |
| Lipid peroxidation | 23 | 1 | 2 | 10 | 10 | 1.122 | 1.429 | 2.551 |
| Protein oxidation | 5 | 0 | 1 | 1 | 3 | 335 | 248 | 583 |
| Homocysteine | 8 | 0 | 2 | 1 | 5 | 680 | 843 | 1523 |
| Iron metabolism | 7 | 0 | 2 | 3 | 2 | 332 | 1.066 | 1.398 |
| Nonvitamin ANTIOX* | 34 | 0 | 9 | 12 | 13 | 2.196 | 3.770 | 5.966 |
| Vitamins A, B, C, D, E* | 11 | 0 | 2 | 4 | 5 | 1.099 | 1.486 | 2.585 |
| Glutathione system* | 14 | 1 | 2 | 3 | 8 | 797 | 789 | 1.586 |

O&NS/ANTIOX: nitro-oxidative stress / antioxidant ratio; O&NS toxicity: combined effects of the neurotoxic O&NS biomarkers;
\*Lowered values of these antioxidant profiles are considered to favor MCI.

### Data search

We searched PubMed/Medline, Web of Science, Scopus, and Google scholar databases from inception until 31 July 2021 for peer-reviewed manuscripts written in English and published in peer-reviewed journals. We first selected English papers in peer-reviewed journals and then manually searched more papers in the reference lists and grey literature and included all studies that were part of previous meta-analyses on O&NS in MCI. Moreover, we also searched papers written in other languages, which are mastered by the authors, namely French, German, Dutch, Thai, Italian, and Spanish. The following keywords were used in the search strategy: "Mild cognitive impairment", "Blood Biomarkers", "Oxidative Stress", "Antioxidants", "Zinc", "Vitamin B", "Albumin", "SOD", "Lipid hydroperoxides", "CAT", "GSH", "Folic", "Coenzyme Q10", "Vitamin E", "Vitamin C", "Vitamin A", " **ESF Table 2** lists all key words used in this study.

**Table 2.** Results of meta-analysis performed on the nitro-oxidative stress (O&S) and antioxidants (ANTIOX) profiles in Mild Cognitive Impairment versus healthy controls.

| Outcome feature sets | n | SMD | 95% CI | z | p | Q | df | p | I<sup>2</sup> (%) | $\tau^2$ | $\tau$ |
| --- | --- | --- | --- | --- | --- | --- | --- | --- | --- | --- | --- |
| O&NS/ANTIOX | 46 | 0.378 | 0.250;0.506 | 5.780 | <0.001 | 323.830 | 45 | <0.001 | 86.104 | 0.149 | 0.386 |
| O&NS | 36 | 0.505 | 0.319;0.691 | 5.333 | <0.001 | 356.534 | 35 | <0.001 | 90.183 | 0.271 | 0.521 |
| O&NS toxicity | 30 | 0.621 | 0.377;0.865 | 4.994 | <0.001 | 363.688 | 29 | <0.001 | 92.026 | 0.402 | 0.634 |
| Lipid peroxidation | 23 | 0.774 | 0.416;1.132 | 4.236 | <0.001 | 355.497 | 22 | <0.001 | 93.811 | 0.686 | 0.828 |
| Protein oxidation | 5 | 0.512 | 0.058;0.967 | 2.210 | 0.027 | 23.952 | 4 | <0.001 | 83.300 | 0.208 | 0.456 |
| Homocysteine | 8 | 0.320 | 0.059;0.581 | 2.404 | 0.016 | 40.922 | 7 | <0.001 | 82.894 | 0.114 | 0.338 |
| Iron metabolism | 7 | 0.325 | -0.071;0.720 | 1.610 | 0.107 | 42.258 | 6 | <0.001 | 85.802 | 0.222 | 0.472 |
| Nonvitamin ANTIOX* | 34 | 0.347 | 0.168;0.527 | 3.799 | <0.001 | 263.291 | 33 | <0.001 | 87.466 | 0.223 | 0.472 |
| Vitamin ANTIOX* | 11 | 0.564 | 0.129;0.999 | 2.543 | 0.011 | 231.360 | 10 | <0.001 | 95.678 | 0.503 | 0.709 |
| Glutathione system* | 14 | 0.725 | 0.269;1.282 | 3.112 | 0.002 | 171.867 | 13 | <0.001 | 92.436 | 0.618 | 0.786 |

SMD: standardized mean difference, 95% CI: 95% confidence intervals
\*Lowered values of these antioxidant profiles are considered to favor mild cognitive impairment

### Eligibility criteria

We included observational case-control, cohort, and randomized controlled trial (RCT) studies which examined the relationship between MCI and O&NS/ANTIOX biomarkers including serum, plasma, erythrocytes, and whole blood cells and which included people aged 63-83 years old, male and female, and met the following inclusion criteria: a) studies reporting on MCI or aMCI as diagnosed using Petersen criteria by clinicians or using neuropsychology test results; b) studies including a healthy or non-MCI control group; and c) studies reporting on O&NS/ANTIOX biomarkers that belong to the predefined profiles as shown in ESF Table 1. We excluded a) studies on O&NS/ANTIOX biomarkers in urine and brain, as well as animal and genetic studies; b) case reports, case series, and cohort studies without a control group, and RCTs without baseline data and controls; c) systematic reviews and meta-analyses; d) studies that did not define MCI as described above; e) studies reporting on comorbidities between MCI and major medical or psychiatric disorders; f) studies that did not report the predefined O&NS/ANTIOX biomarkers as shown in ESF Table 1; g) studies presenting duplicated data; and h) studies not reporting mean and standard deviation (SD) or standard error (SE) values of the biomarkers. The authors of the latter studies were contacted and asked to supply means, standard deviations, and the number of cases and controls. We estimated the mean and SD using the formulas provided by Luo et al. (2018) and Wan et al. (2014) or employed the Webplotdigitizer platform (Rohatgi, 2021) to estimate mean and SD values from graphs. We documented all reasons for excluding studies from the analysis.

### Screening and data extraction

The first author (GN) performed the initial screening, extracting the titles and abstracts of relevant papers to determine eligibility before extracting the full text of papers. GN then extracted the critical data from the full-text versions and entered it into a predefined Excel spreadsheet. If the first author (GN) finished compiling all the information, the second author (AV) double-checked all the data in the spreadsheet. In the event of a disagreement, the last author (MM) was consulted. Data from all eligible studies were entered into the predefined Excel spreadsheet template, including author names, publication years, O&NS/ANTIOX profiles and biomarker types, mean concentration values with SD and sample sizes, MCI participants, and healthy control participants. From each study, we extracted the following data: the design and setting, the mean age, gender and ethnic distribution, comorbidities, the neurocognitive tests used, and criteria for MCI assessment, medium (e.g., plasma or serum), study latitude, and methodological quality scores (quality and redpoint checklists) as shown in **ESF Table 3**. We modified and adapted the confounder’s and red point scales used in previous studies by this lab (Andrés-Rodríguez et al., 2020; Vasupanrajit et al., 2021a; Vasupanrajit et al., 2021b) for O&NS research. The first scale evaluates the methodological quality of the O&NS/ANTIOX studies, focusing on the more critical aspects such as sample size, confounder control, age and gender matching, medical comorbidities, and so on. The methodological quality is graded from 0 to 10, with lower scores indicating lower quality and higher scores indicating higher quality. The redpoint score (see ESF, Table 3) can be used to determine the extent to which important confounders were overlooked, introducing bias into the O&NS/ANTIOX studies. When all confounding variables were included, the total score was set to 0, and when no control was present, the total score was set to 25.

**Table 3.** Publication bias of the nitro-oxidative stress (O&NS) and antioxidant (ANTIOX) profiles in Mild Cognitive Impairment

| Outcome | Fail safe n | Z Kendall's $\tau$ | P (1-tailed) | Egger's (df) | P (1-tailed) | Missing studies (side) | Adjusted SMD |
| --- | --- | --- | --- | --- | --- | --- | --- |
| O&NS/ANTIOX | 2380 | 1.950 | 0.025 | 1.893 (44) | 0.032 | 7 (Right) | 0.471 (0.340;0.602) |
| O&NS | 2223 | 1.430 | 0.067 | 0.985 (34) | 0.166 | 8 (Right) | 0.704 (0.509;0.898) |
| O&NS toxicity | 1732 | 2.177 | 0.015 | 1.418 (28) | 0.084 | 5 (Right) | 0.812 (0.554;1.069) |
| Lipid peroxidation | 1121 | 2.324 | 0.010 | 1.295 (21) | 0.105 | 1 (Right) | 0.857 (0.489;1.228) |
| Protein oxidation | 23 | 1.225 | 0.110 | 2.524 (3) | 0.043 | 2 (Left) | 0.256 (-0.166;0.679) |
| Homocysteine | 70 | 0.123 | 0.450 | 0.593 (6) | 0.287 | 0 | 0.319 (0.059;0.580) |
| Iron metabolism | 22 | 0.000 | 0.500 | 0.601 (5) | 0.287 | 0 | 0.325 (-0.071;0.720) |
| Nonvitamin ANTIOX* | 939 | 2.253 | 0.012 | 0.850 (32) | 0.200 | 6 (Right) | 0.470 (0.295;0.645) |
| Vitamin ANTIOX* | 252 | 0.778 | 0.218 | 1.585 (9) | 0.073 | 4 (Right) | 0.889 (0.405;1.373) |
| Glutathione system* | 257 | 1.861 | 0.031 | 2.358 (12) | 0.018 | 3 (Right) | 1.268 (0.694;1.841) |

\*Lowered values of these antioxidant profiles are considered to favor mild cognitive impairment

### Primary and secondary outcomes

The primary outcome was the O&NS/ANTIOX profile in MCI participants *versus* healthy controls. In order to delineate which components of the O&NS/ANTIOX are most relevant to MCI, we performed secondary outcomes analyses on O&NS, O&NS-toxicity, lipid-peroxidation, iron-metabolism, homocysteine, protein-oxidation, nonvitamin-ANTIOX, vitamins (A, B, C, D, E), and glutathione profiles (see ESF table 1). To delineate which solitary biomarkers contributed most to the alterations in those specific O&NS and ANTIOX profiles, we performed additional meta-analyses on the single biomarkers when there were at least two studies.

### Data analysis

The Comprehensive Meta-Analysis V3 software was used to conduct meta-analysis and meta-regression analysis. The nine synthetic scores were compared between MCI participants and healthy controls by calculating the mean values of the outcomes in a specific profile while assuming dependence. The direction of the O&NS variables was determined to be positive (increasing levels for MCI), whereas the direction of antioxidant biomarkers was determined to be negative (thus, lowered levels for MCI). We conducted the analysis using a random-effects model with restricted maximum-likelihood on the assumption that the characteristics of the several studies varied. We calculated the standardized mean difference (SMD) using 95% confidence intervals (CI). A significance level of 0.05 indicates statistically significant pooled effect sizes. Heterogeneity was delineated as explained previously using tau-squared values (Vasupanrajit et al., 2021a), although we also calculated Q and I^2^ metrics. We performed sensitivity analyses using the leave-one-out method to check the robustness of the effect sizes and between-study heterogeneity. To assess small study effects such as publication bias, we used Egger’s regression intercept (using one-tailed p-values), Kendall tau with continuity correction (using one-tailed p-values), and the classic fail-safe N method. Additionally, we used funnel plots (study precision *versus* SMD), which simultaneously display the observed and imputed missing values, to detect small study effects.

## Results

### Search results

**Figure 1** shows the PRISMA flow diagram and gives information about the different phases of this review, the results of our search with the number of records identified, excluded, and included. We screened 2.352 records after removing 188 studies. Forty-nine articles were assessed in the systematic review and three of those studies could not be included in the MA for reasons shown in **ESF Table 4**. As a result, the meta-analysis was performed on 46 studies (Al-Rawaf et al., 2021; Arce-Varas et al., 2017; Ayromlou et al., 2018; Balmus et al., 2017; Bermejo et al., 2008; Boccardi et al., 2021; Cervellati et al., 2013; Cervellati et al., 2014a; Cervellati et al., 2014b; Chico et al., 2013; Chmatalova et al., 2017; Dong et al., 2008; Du et al., 2019; Gironi et al., 2011; Gironi et al., 2015; Guidi et al., 2006; Gunduztepe et al., 2020; He et al., 2016; Irizarry et al., 2007; Iuliano et al., 2010; Kim et al., 2013; Lanyau-Domínguez et al., 2020; Lee et al., 2013; Liu et al., 2016; Lopez et al., 2013; Ma et al., 2017; Martin-Aragon et al., 2009; Martinez de Toda et al., 2019; McFarlane et al., 2020; Mota et al., 2015; Mueller et al., 2012; Mufson and Leurgans, 2010; Negahdar et al., 2015; Padurariu et al., 2010; Perrotte et al., 2019; Praticò et al., 2002; Rembach et al., 2013; Rinaldi et al., 2003; Rita Cardoso et al., 2014; Sirin et al., 2015; Squitti et al., 2011; Sultana et al., 2013; Torres et al., 2011; Ulstein and Bohmer, 2017; Yuan et al., 2016; Zhou et al., 2020). **ESF Table 5** summarizes the results of all studies included in the meta-analysis.

**Figure 1.**
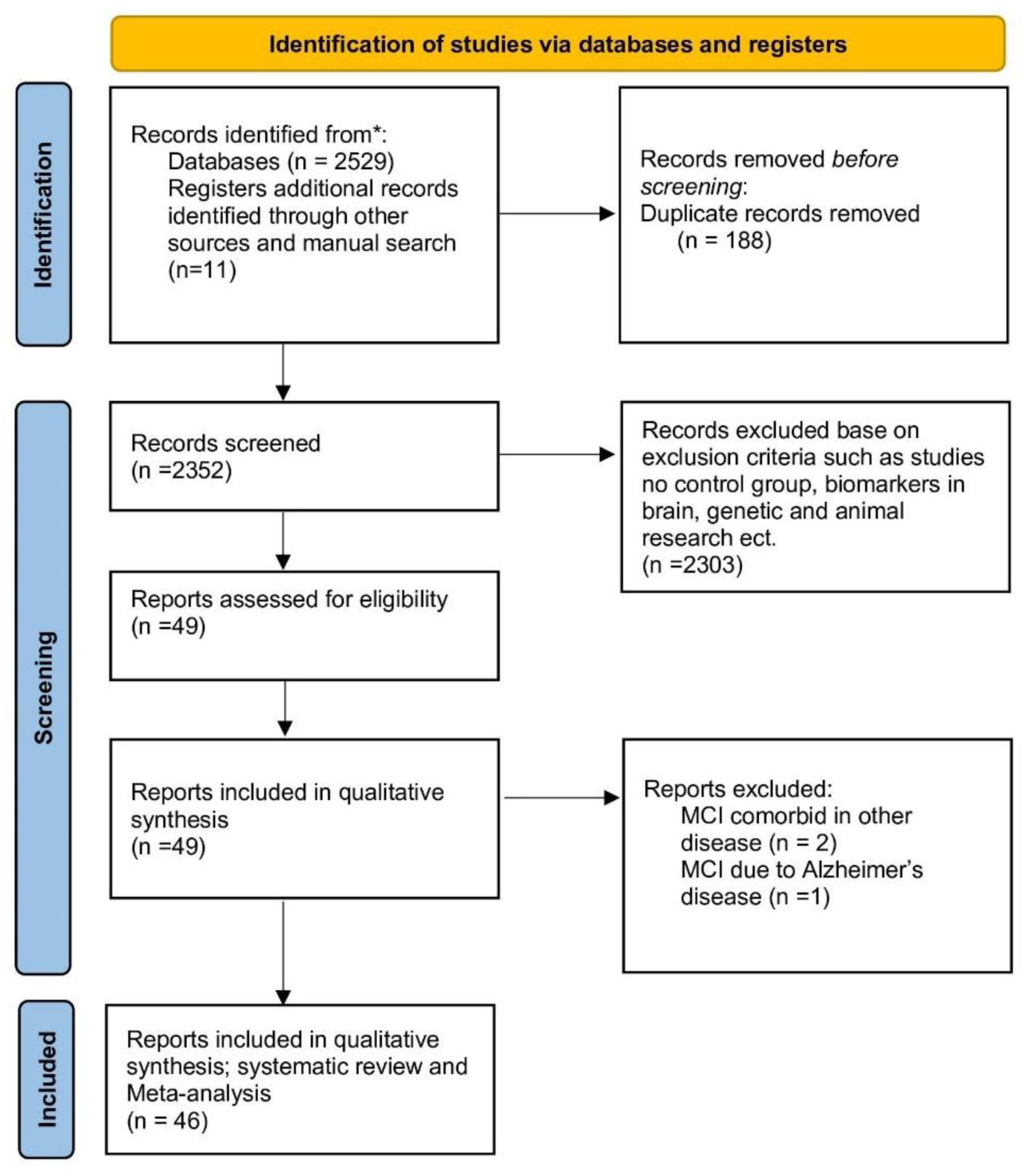
Prisma 2020 diagram

The MA included participants from all continents, namely 1 study from Saudi Arabia, the Czech Republic, South Korea, Cuba, Poland, Portugal, Malaysia, Canada, Norway, Australia, Germany, and Canada; 2 studies from Turkey, Romania, Brazil, and Iran; 5 studies from USA and Spain; 6 studies from China, and 10 studies from Italy. The meta-analysis comprised 36 case-control studies, 3 cross-sectional studies, 5 cohort studies, 1 retrospective study and 1 study with a longitudinal design. Almost all studies reported plasma and serum assays with 8 studies reporting combined erythrocyte, hemoglobin, and mitochondria media.

### The primary outcome O&NS/ANTIOX

**Table 1** displays the overall meta-analysis data of the 46 studies performed on the O&NS/ANTIOX profiles with a total participant sample of 9.861 including 3.798 MCI cases and 6.063 healthy controls. ESF table 4 shows that the meta-analysis excluded three studies due to comorbidity issues in the MCI and control groups (Rozzini et al., 2018; Vinothkumar et al., 2017; Zheng et al., 2019). Rozzini et al. (2018) found that free and total copper were higher in MCI due to AD than in healthy controls. Vinothkumar et al. (2017) reported reduced plasma SOD, CAT, GSH peroxidase (Gpx) in patients with chronic kidney disease (CKD) and cognitive dysfunction when compared to patients with CKD without cognitive dysfunction. Zheng et al. (2019) found lower folate levels in type 2 diabetes with MCI (T2D-MCI) when compared to controls, whereas there were no significant differences in HCL-c.

Table 1 shows that 18 studies had confidence intervals that were entirely positive of zero, 22 studies had confidence intervals that overlapped zero but had SMD values greater than zero, and 6 studies had confidence intervals that overlapped zero but had a mean lower than zero. There were no studies that reported confidence intervals that were entirely negative of zero. **Table 2** shows the results of a random-effect meta-analysis on the 46 studies and indicates that the O&NS/ANTIOX profile was significantly higher in MCI participants than in healthy controls, with a moderate effect size of 0.378. The forest plot of the O&NS/ANTIOX markers in MCI *versus* controls is shown in **Figure 2**. We performed a sensitivity analysis after eliminating two studies with higher SMD values (> 3) (Balmus et al., 2017; Padurariu et al., 2010) and found that the effect size was still significant (SMD=0.335, 95%; CI: 0.218; 0.452, p<0.0001, tau-squared=0.121). **Table 3** shows that there was some publication bias, with seven studies missing on the right side of the funnel plot yielding an adjusted point estimate of 0.471 (95% CI: 0.340; 0.602).

**Figure 2.**
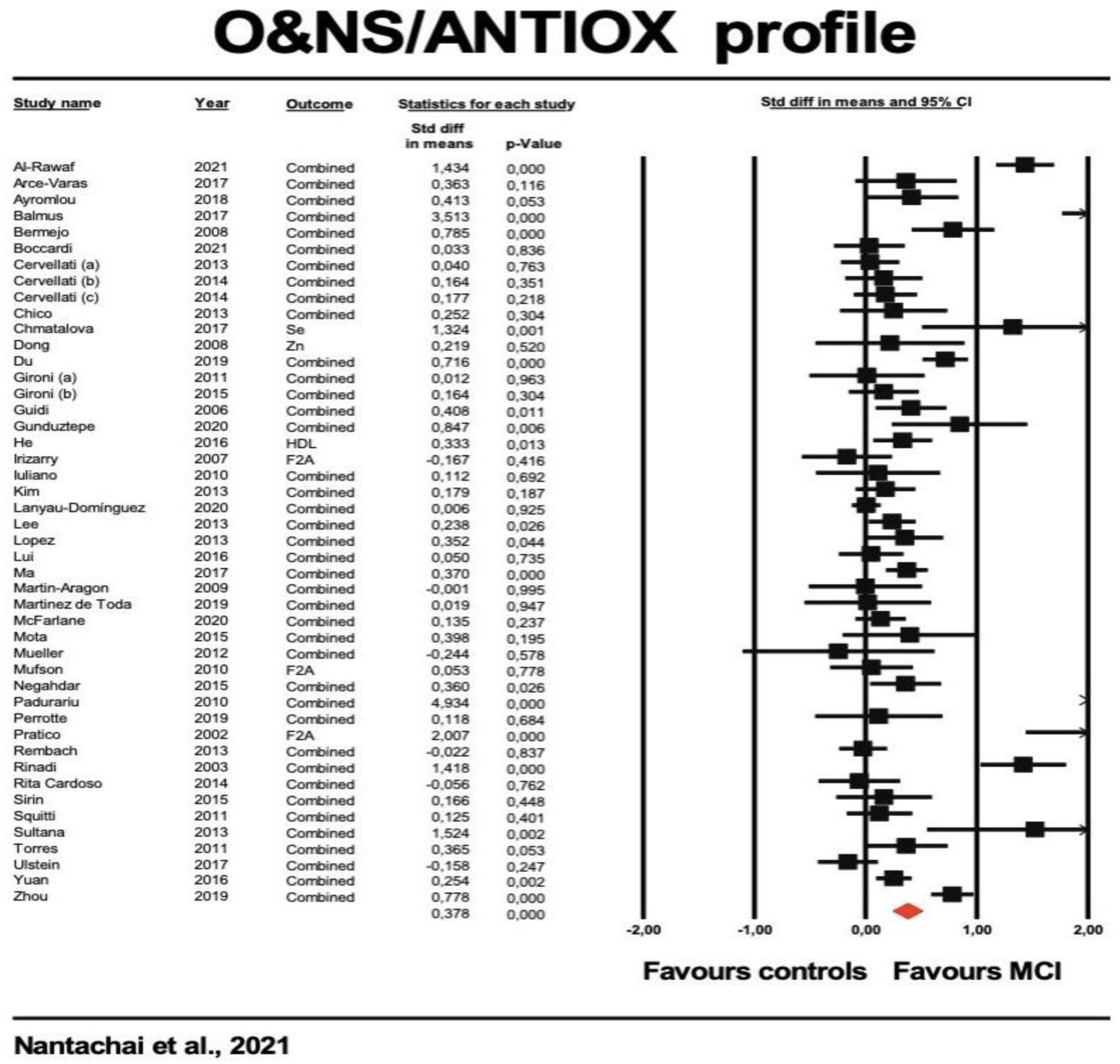
Forest plot with results of meta-analysis performed on 46 studies reporting on the nitro-oxidative stress / antioxidant (O&NS/ANTIOX) ratio in Mild Cognitive Impairment (MCI) versus controls.

Group analysis showed that there were no significant differences between serum, plasma, and other medium (p=0.121) and between case-control studies and other study designs (p=0.354). Meta-regression showed a significant effect of the mean age of the MCI group (z=-2.38, p=0.017) and that there were no significant effects of sex distribution (p=0.841), Caucasian/non-Caucasian ratio (p=0.060), and latitude (p=0.794). ESF Table 3 shows that the median quality score was 4 (min=2.5, max=6), and the median redpoint score 16 (min=8.25, max=21.5). Meta-regression did not reveal significant effects of the quality (p=0.217), and redpoint (p=0.484) scores.

### The secondary outcome profiles O&NS and O&NS-toxicity profiles

There were 36 studies including the O&NS profile reporting on 2.325 MCI cases *versus* 4.257 healthy controls and 30 studies on O&NS-toxicity reporting on 1.795 MCI participants and 2.275 healthy controls. Table 1 shows that 13 O&NS studies had confidence intervals which were entirely on the positive side of zero, while only one study showed a confidence interval which was entirely on the negative side of zero, and 18 studies showed overlapping intervals, 15 with SMD greater than zero and 3 with SMD values lower than zero. Twelve O&NS-toxicity profile studies showed confidence intervals that were entirely on the positive side of zero, and only 1 study showed a confidence interval that was entirely on the negative side of zero. Seventeen studies showed overlapping confidence intervals, namely 13 studies with SMD greater than zero and 4 studies with SMD values lower than zero. ESF Table 4. displays the three studies that were excluded from the MA but were included in the systematic review.

Table 2 shows that both the O&NS and O&NS-toxicity profiles were higher in MCI than in controls with medium to high effect sizes, namely 0.505 (CI: 0.319; 0.691) and 0.621 (CI: 0.377; 0.865), respectively. **Table 3** shows that there was some publication bias, with eight and five studies missing on the right side of the funnel plot of O&NS and O&NS toxicity, respectively. One of the toxic O&NS products, namely copper, showed a small but significant effect size for MCI (SMD=0.197, CI: 0.052; 0.341, p=0.007, n=5).

### Lipid-peroxidation, protein-oxidation, homocysteine, and iron profiles

Table 1 shows that the 23 studies on lipid-peroxidation included 1.122 MCI subjects *versus* 1.429 healthy controls. The 5 studies on the protein-oxidation profile included 335 MCI subjects *versus* 248 healthy controls and the 8 studies on the homocysteine profile reported on 680 MCI subjects *versus* 843 healthy controls. The 7 studies on the iron metabolism profile reported data on 332 MCI *versus* 1.066 healthy controls. Table 1 shows the distribution of the confidence intervals of these 4 profiles with respect to the zero SMD values. Ten lipid peroxidation studies (see **Figure 3**) showed confidence intervals which were on the positive side of zero, while only one study showed confidence intervals that were entirely on the negative side of zero; 10 studies had confidence intervals that overlapped zero but had SMD values greater than zero, and 2 studies had confidence intervals that overlapped zero but had a mean lower than zero.

**Figure 3.**
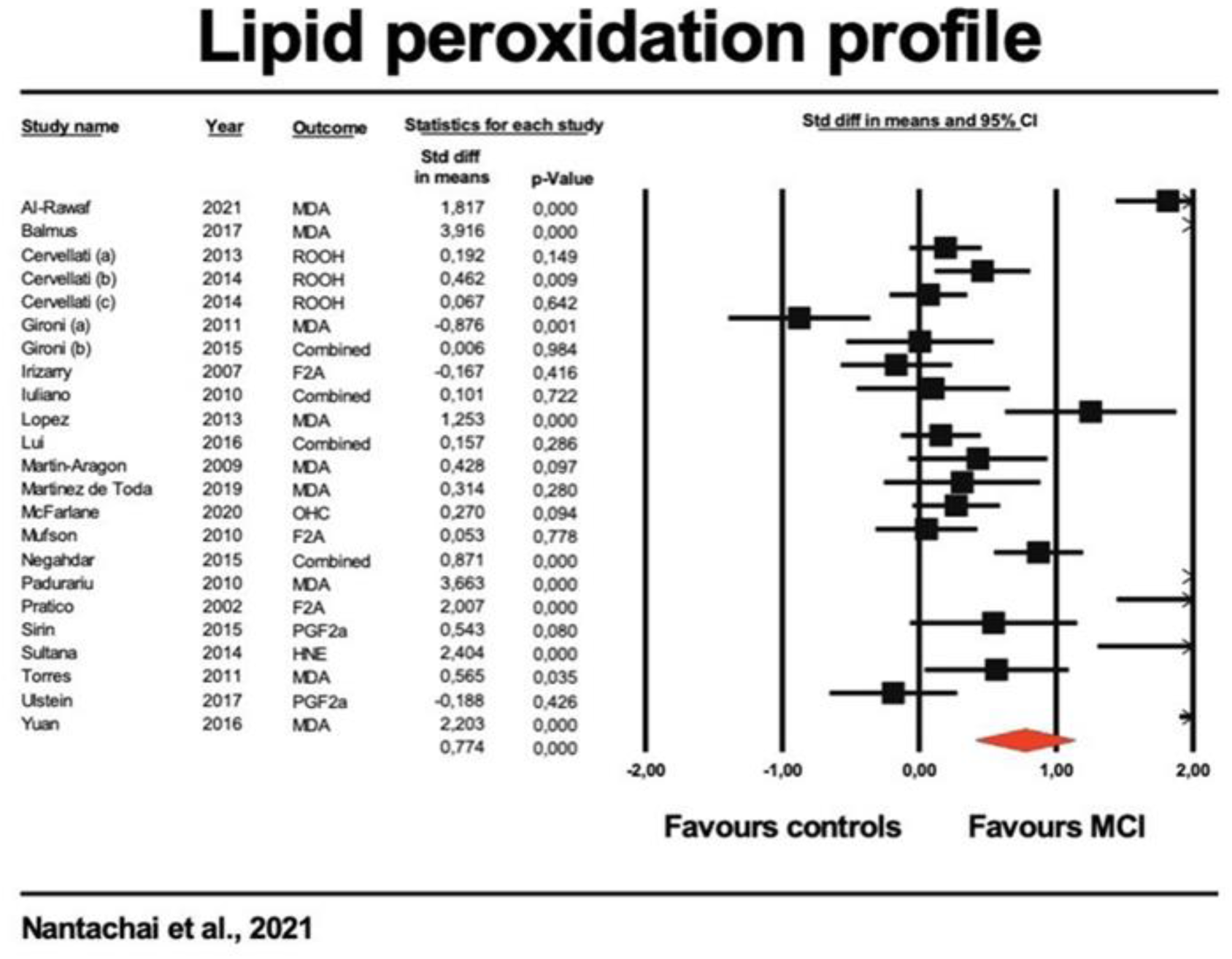
Forest plot with results of meta-analysis performed on lipid peroxidation profile in Mild Cognitive Impairment (MCI) versus controls.

Table 1 shows the confidence intervals of the protein oxidation profile and that 3 studies had confidence intervals that were entirely positive of zero, 1 study had confidence intervals that overlapped zero but had SMD values greater than zero, 2 studies had confidence intervals that overlapped zero of which one had a mean higher than zero and one a mean lower than zero. Table 1 shows that 5 homocysteine studies had confidence intervals which were entirely on the positive side of zero, while 0 studies showed confidence intervals which was entirely on the negative side of zero, 3 studies showed overlapping intervals, 1 with a mean greater than zero and 2 with mean values lower than zero.

Table 2 shows that the lipid peroxidation, protein oxidation and homocysteine, but not iron, profiles were significantly higher in MCI subjects than in healthy controls. Lipid peroxidation showed the highest effects size, namely 0.774 (0.416; 1.132). A meta-analysis performed on the 11 studies which examined MDA/TBARS data showed a very high effect size, namely SMD of 1.222 (CI: 0.574; 1.870, p<0.001, n=11, tau-squared=1.094). Meta-analyses performed on the two carbonyl and three AOPP studies showed a high effect size for carbonyls (SMD=1.038, CI: 0.581; 1.496, p<0.001, tau-squared is <0.001), while the AOPP analysis yielded non-significant results (SMD=0.253, CI: -0.207; 0.713, p=0.281, tau-squared = 0.137).

Table 3 shows that there was some publication bias in the lipid peroxidation and protein-oxidation, but not homocysteine and iron, profile data. One study was missing on the right side of the funnel plot of the lipid peroxidation plot with an adjusted point estimate of 0.857 (CI: 0.489; 1.228). For protein-oxidation, two studies were missing on the left site and the point estimate was no longer significant, namely 0.256 (CI: -0.166; 0.679).

### The antioxidant profiles

Three different biomarker profiles considered ANTIOX outcomes, namely 34 studies reporting on the nonvitamin-ANTIOX profile (2.196 MCI *versus* 3.770 healthy controls), 11 studies on vitamins A, B, C, D, E (1.099 MCI *versus* 1.486 healthy controls), and l4 studies on the glutathione profile (797 MCI *versus* 789 healthy controls) (see table 1). Table 1 shows that 13 non-vitamin ANTIOX studies had confidence intervals which were entirely on the positive side of zero, while no studies showing confidence intervals which were entirely on the negative side of zero, 21 studies showed overlapping intervals with 12 studies showing a mean greater than zero and 9 with mean values lower than zero. Five vitamin ANTIOX studies had confidence intervals which were entirely on the positive side of zero, while no studies showed confidence intervals which were entirely on the negative side of zero, 6 studies showed overlapping intervals with 4 studies showing a mean greater than zero and 2 with mean values lower than zero. Eight glutathione profile studies had confidence intervals which were entirely on the positive side of zero, while one study showed confidence intervals which were entirely on the negative side of zero, 5 studies showed overlapping intervals with 3 study showing a mean greater than zero and 2 with a mean value lower than zero.

Table 2 shows that the 3 ANTIOX biomarker profiles were significantly lower in MCI than in controls and that the glutathione system showed the greatest effect size, namely 0.725 (0.269; 1.182). These three profiles showed some bias with a few missing studies on the right site. The adjusted SMD for glutathione was 1.268 (0.694; 1.841). A secondary analysis showed that Gpx was significantly reduced in MCI with and effect size of 0.948 (CI: 0.175; 1.721, p=0.016, n=9, tau-squared=1.135). We performed a sensitivity analysis after eliminating two studies with very high SMD values (Balmus et al., 2017; Padurariu et al., 2010) and found that the effect size was still significant but reduced to 0.390 (CI: 0.001; 0.779, p=0.050, n=5, tau-squared=0.137).

No significant differences could be detected in glutathione reductase (p=0.560, n=4), CAT (p=0.433, n=5), HDL-c (p=0.168, n=10), and SOD (p=0.167, n=9). A meta-analysis performed on all antioxidant indices combined (BAP, FRAP, RAP, TAC, PAO) showed significantly lowered levels in MCI than in controls with a high effect size (SMD=0.759, CI: 0.197; 1.322, p=0.008, n=9, p=0.008, tau-squared=0.714). Another meta-analysis conducted on the combined SOD, albumin, selenium, transferrin, HDL-c, thiol groups, and zinc data showed that this synthetic score was significantly lower in MCI than in controls (SDM=0.341, CI: 0.098; 0.530, p=0.004, n=26, tau-squared=0.262). However, none of the solitary biomarkers was significantly decreased in MCI. Folate levels were significantly lower in MCI than in controls although the effect size (0.179) was marginal (CI: 0.000; 0.358, p=0.050, n=6, p=0.050, tau-squared=0.028). Vitamin A (SDM=0.442; CI: 0.157; 0.726, p=0.002, n=6, p=0.002, tau-squared=0.094) and vitamin C (SDM=0.608; CI: 0.094; 1.123, p=0.021, n=5, p=0.021, tau-squared=0.303) levels were significantly lower in MCI than controls with a medium effect size. Nevertheless, vitamin E (p=0.152, n=5) and vitamin B12 (p=0.118, n=6) were not significantly different between both study groups.

## Discussion

### O&NS biomarkers in MCI

The first major findings of our study are that a) the meta-analysis revealed that the O&NS/ANTIOX ratio was significantly higher in MCI participants when compared to controls, and b) the systematic review revealed that 40 studies reported O&NS/ANTIOX mean values favoring MCI, while only six studies reported a mean not favoring MCI. These findings suggest that O&NS processes and/or lowered antioxidant defenses are associated with MCI. As such, our results extent those of previous meta-analysis studies which showed increases in individual O&NS and antioxidant biomarkers in aMCI as compared with controls (Ashley et al., 2019; Schrag et al., 2013). Moreover, our systematic review and meta-analysis was able to delineate the specific O&NS and ANTIOX profiles that contributed to the increased O&NS/ANTIOX ratio in MCI (discussed below).

The second major finding is that lipid-peroxidation is significantly increased in MCI as compared with controls with a large effect size of 0.774 SMD. The biomarkers included in this profile were MDA or TBARS, ROOH (hydroperoxides), LOOH (lipid hydroperoxides), OH-cholesterol, and oxidized LDL. As such, our results show that an upregulation of ROS, peroxides, lipid peroxides, and consequent aldehyde formation are hallmarks of MCI. Moreover, one of the final and most detrimental outcomes of lipid peroxidation, namely aldehyde formation, showed a very high effect size (SMD=1.222; CI: 0.574; 1.870) for MCI versus controls. These results extend those of previous reports which suggested that MDA/TRARS levels may be useful biomarkers to help diagnosing MCI (Kandlur et al., 2020) and those of a previous meta-analysis that plasma TBARS (but not erythrocyte TBARS) has a large effect size (0.79, CI: 0.41; 1.16) for MCI (Schrag et al., 2013). Aldehydes are primarily produced as a byproduct of hydroxyl radicals and increased peroxynitrite production during the lipid hydroperoxide chain reactions (Maes et al., 2018). O&NS and especially aldehyde formation play a key role in neuronal death in neuroinflammatory and neurodegenerative disorders including Alzheimer’s and Parkinson’s disease, and multiple sclerosis (Sayre et al., 2008). Moreover, increased MDA levels are associated with ageing and neurocognitive impairments (Negahdar et al., 2015).

It is interesting to note that in our meta-analysis, lipid peroxidation was more strongly associated with MCI that protein-oxidation biomarkers. The latter profile comprised AOPP and protein carbonyls, with the latter showing a high effect size (1.038, CI: 0.581; 1.496), while a previous meta-analysis performed on AOPP data yielded non-significant results (SDM=0.253, CI:

-0.207; 0.713). One previous study suggested that using protein carbonyls markers may be helpful for diagnosing MCI (Kandlur et al., 2020), while Schrag et al. (2013) observed no significant differences in protein carbonyls between both Alzheimer’s disease and MCI and controls. However, after adjusting for possible bias in the protein oxidation data all significance disappeared, although both AOPPP and carbonyl data contribute to the increased O&NS-associated neurotoxic profile in MCI.

Increased AOPP formation occurs as a result of increased hypochlorous acid stress caused by increased myeloperoxidase activity in conjunction with increased peroxide and peroxynitrite production caused by SOD (Huang et al., 2013; Maes et al., 2018). Chlorinative stress may contribute to apoptosis, damage to DNA, tissue lesions and inflammation and plays a role in neurodegenerative disease (Casciaro et al., 2017). Protein carbonyl levels, on the other hand, are another type of protein oxidation products as a consequence of increased ROS with formation of high molecular weight carbonyls (keto, aldehyde, or lactam groups) in proteins, phospholipids, or low molecular weight carbonyls resulting from oxidative cleavage reactions (Fedorova et al., 2014).The resulting carbonyl stress may result in cell toxicity, apoptotic cell death, inflammation, and increased immunogenicity (Fedorova et al., 2014).

The homocysteine profile was only marginally associated with MCI with a low effect size of 0.241 (CI: 0.042; 0.440). Homocysteine is a sulfur-containing amino acid that plays a significant role in methionine metabolism and increased homocysteine levels contribute to ROS production (Faraci and Lentz, 2004). Moreover, increased homocysteine may damage neuronal DNA and promote excitotoxicity via stimulation of N-methyl-D-aspartate receptors thereby causing apoptosis and cell death (Diaz-Arrastia, 2000; Ho et al., 2002; Reynolds, 2006).

No significant differences could be found in the iron profile between MCI individuals and healthy controls. One study reported that an increase in serum copper / iron ratio predicted the transition of MCI to Alzheimer’s disease (Mueller et al., 2012). Thus, it may be that the increased levels of copper may be more important than the smaller decreases in serum iron. This is further substantiated in the current meta-analysis which showed significantly higher copper values in MCI than in controls, albeit with a small effect size. The possible contribution of free copper to MCI were also observed by Squitti et al. (2011). Most importantly, the results indicate that increases in iron concentrations, which may be another source of O&NS, are not associated with MCI.

### Antioxidant levels in MCI

The third major finding of this study is that the ANTIOX profiles, including the glutathione system (GSH, GST peroxidase, and GSH reductase) and non-vitamin and vitamin antioxidant defenses, were significantly reduced in MCI as compared with controls and that the effects size of the glutathione system was as large. In a secondary analysis, we discovered that MCI cases showed significantly lower levels of Gpx and GSH, but not glutathione reductase. Previously, no evidence of a significant decrease in Gpx (serum, plasma, or erythrocytes) was found in MCI or Alzheimer’s disease (Schrag et al., 2013). Furthermore, the latter meta-analysis found no evidence of reduced glutathione reductase levels in Alzheimer’s disease patients. However, another study found that in vivo GSH levels were lower in the hippocampus and frontal cortex of subjects with MCI and Alzheimer’s disease, and that these levels were directly related to cognitive impairments (Mandal et al., 2015). Furthermore, GSH reductions distinguished MCI from controls with high accuracy (sensitivity: 87.5 percent and specificity: 100 percent).

The glutathione system is an important player in maintaining redox balance in intracellular redox pathways, particularly in the brain, explaining why decreased GSH defenses with increased ROS can have disastrous consequences, including oxidative CNS damage (Bermejo et al., 2008; Pocernich and Butterfield, 2012). Furthermore, GSH is required for xenobiotic detoxification, and low GSH levels are linked to neuronal apoptosis and mitochondrial dysfunction, whereas GSH-dependent N-methyl-D-aspartate receptor modulation regulates glutamate excitotoxicity (Bjørklund et al., 2021; Morris et al., 2014). Furthermore, decreased GSH defenses and GSH oxidation are linked to dysfunctions in macrophage functions, dendritic cell maturation and differentiation, antigen presentation, neutrophil respiratory burst and extracellular trap production, and T cell regulatory functions (Morris and Maes, 2021) . It is important to note in this regard that MCI is frequently associated with activated immune-inflammatory pathways (Saleem et al., 2015; Trollor et al., 2010; Zheng et al., 2019). The current meta-analysis also discovered that the multivitamin profile, which included vitamins A, B (B1, B2, B6, B9, and B12), C, D, and E, was significantly lower in MCI people compared to controls, and that vitamins A, B9, and C, in particular, contributed to these effects.

Previous research suggested that low levels of vitamins A, B, C, D, and E, as well as beta-carotene, may favor MCI (Ayromlou et al., 2018). A previous meta-analysis found that serum α-tocopherol levels were not significantly altered in subjects with MCI, despite the fact that serum α-tocopherol levels were significantly reduced by about 20% in Alzheimer’s disease (Schrag et al., 2013). According to the same meta-analysis, vitamin A levels in two MCI studies were not altered, whereas vitamin A levels in Alzheimer’s disease patients were reduced by 25%. Furthermore, both β- and α-carotene levels were reduced in Alzheimer’s disease, particularly α-carotene (Schrag et al., 2013). Folate and vitamin B12 levels were lower in MCI than in controls (Ma et al., 2017), and low folate levels were associated with cognitive impairments (Kim et al., 2013). This is significant because folate levels may protect against rising homocysteine levels and thus homocysteine-induced toxicity (Ma et al., 2017). Adequate folate intake at baseline is associated with improved cognitive reserve, whereas low B6, B9, and B12 intake may be associated with neurocognitive dysfunctions via effects on methylation of specific genes (An et al., 2019).

The non-vitamin antioxidant profile, which included antioxidants such as SOD, albumin, selenium, transferrin, HDL-c, thiols, and zinc, was also significantly reduced in MCI with a medium effect size. It is worth noting that in Schrag et al. (2013)’s meta-analysis, SOD and CAT levels were not significantly associated with Alzheimer’s disease, whereas total antioxidant capacity was significantly reduced in Alzheimer’s disease with an effect size of -0.85 (CI: 1.29 to -0.41). Transferrin and ceruloplasmin levels did not change in Alzheimer’s disease, but ceruloplasmin enzyme activity did (Schrag et al., 2013). The latter systematic review reported that total antioxidant capacity was significantly reduced in Alzheimer’s disease patients.

### Why assess and analyze biomarker profiles

It is worth noting that, while none of the individual non-vitamin ANTIOX biomarkers (SOD, albumin, selenium, transferrin, HDL-c, thiol groups, and zinc) were reduced in MCI, the overall profile was significantly reduced with a medium effect size. This suggests that these antioxidants contribute to overall reductions in antioxidant defenses, even though such effects may not be detected when looking at solitary antioxidants. TAC, BAP, FRAP, PAO and RAP indices of a more general antioxidant capacity of plasma/serum were also significantly lower in people with MCI compared to controls. As a result, MCI is characterized by a significant decrease in overall antioxidant defenses, with deficiencies in the GSH system being the principal contributor. It should be noted that we used synthesized scores of different biomarkers belonging to a specific O&NS domain in the current meta-analysis. As previously stated, this is a far more precise and powerful approach than performing a meta-analysis on individual biomarkers (Vasupanrajit et al., 2021a; Vasupanrajit et al., 2021b). For example, if a study reports on the analysis of two lipid peroxidation biomarkers (for example, LOOH and MDA) in the same patients and controls, LOOH and MDA could be analyzed separately in two separate meta-analyses. This approach, however, is questionable because these measurements are not independent because they were performed on the same population and follow the same pathway, namely lipid peroxidation with subsequent aldehyde formation. Furthermore, if one of these biomarkers is significantly elevated in MCI while the other is not, a meta-analysis based on a single biomarker may yield incorrect conclusions about lipid peroxidation. As a result, conducting two separate meta-analyses on LOOH and MDA (both of which reflect lipid peroxidation) or a single meta-analysis on a single biomarker when more biomarkers are available precludes generalization of the findings to lipid peroxidation.

In our meta-analysis, on the other hand, we averaged biomarkers from the same profile to create a synthetic score and then averaged those scores across all studies. By doing so, we were able to generate robust composite indices for a wide range of functionally meaningful profiles using all available data and without making any selections. As a result, all available data is incorporated into composite profile scores, which serve as meaningful indices of the O&NS profiles and are thus generalizable to MCI.

### Limitations

To begin, our meta-analysis results would have been more interesting if we could have included more studies on protein oxidation (carbonyls as well as AOPP), the glutathione system (GSH and GSH reductase), zinc, selenium, and vitamins A, B6, E, and D. Furthermore, there have been very few studies that have assessed nitration and nitrosative stress, including measurements of reactive nitrogen species, nitric oxide, nitrotyrosine, and nitrosylation indicators (Morris et al., 2017; Morris et al., 2018), so we were unable to include a specific nitrosative stress profile in our analysis. Finally, a few key O&NS and antioxidant biomarkers, such as myeloperoxidase, coenzyme Q10, and paraoxonase 1, were largely absent. Future research should concentrate on biomarker assays and measure these and other biomarkers in the same well-powered study. Second, this meta-analysis study was unable to include assessments of MCI severity, including neurocognitive impairment severity, or MCI subtypes such as aMCI. Third, the quality of the studies included in our analysis is rather low, and future research on O&NS in MCI should strive for higher scores on our confounder and red point scores, which are used to assess the quality of the included O&NS biomarker studies.

### Conclusions

The study’s main findings are that the ONS/ANTIOX ratio is significantly higher in MCI compared to controls, and that lipid peroxidation with increased aldehyde formation, increased homocysteine, carbonyl levels, and copper levels, and decreased GSH and GSH peroxidase, non-vitamin and vitamin (especially lowered vitamin B9, vitamin A, and C levels) antioxidants contribute to MCI. These findings suggest that increased O&NS and decreased antioxidant defenses are important factors in the development of MCI. It is worth noting that aging is associated with increased O&NS and decreased antioxidant defenses (Guidi et al., 2006; Kandlur et al., 2020), and that these aging-processes in redox regulation may explain, at least in part, the onset of MCI in the elderly. Furthermore, such redox regulation dysfunctions may contribute to the dysregulation and activation of immune-inflammatory pathways, which are also linked to MCI (Morris and Maes, 2021; Saleem et al., 2015).

Our findings indicate that lipid peroxidation, particularly increased aldehyde formation, as well as decreased GSH defenses, vitamin A and C levels, and the homocysteine / folic acid ratio, are drug targets in MCI that should be targeted to improve the redox balance. Treatment with specific antioxidants has been shown to improve cognitive deficits in people with MCI, Alzheimer’s disease, and other neurodegenerative disorders. Treatment with N-Acetyl-L-Cysteine (NAC), an antioxidant that activates the GSH system and has immune-regulatory properties, has shown some clinical efficacy in clinical trials in people with MCI, Alzheimer’s disease, and Parkinson’s disease, particularly when combined with conventional therapies for these disorders (Tardiolo et al., 2018). Elevations in GSH via NAC and glutamylcysteine ethyl ester (GCEE) administration may improve neuroprotection in the brain, thereby improving cognitive impairments and possibly preventing the progression of MCI to Alzheimer’s disease (Pocernich and Butterfield, 2012). Vitamin B supplements including folate appear to be effective in the prevention of cognitive decline in elderly people (Li et al., 2021). Moreover, folate administration may lower homocysteine levels and O&NS (Zhou and Chen, 2019) and folate administration, as well as the combination of folate with other nutrients, may be beneficial in the treatment of MCI and Alzheimer’s disease (Puga et al., 2021). Overall, measurements of O&NS (MDA and homocysteine) and antioxidants (folate and the GSH system, vitamins A and C) should be performed in subjects with MCI, and treatment with GSH-promoting supplements (e.g., NAC), as well as vitamin A, B and C, may improve the redox imbalance and, most likely, MCI.

## Availability of data and materials

The dataset (CMA file) generated during and/or analyzed during the current study will be available from MM upon reasonable request and once the dataset has been fully exploited by the authors

## Author contributions

Conceptualization, M.M. and G.N., formal analysis, M.M. and G.N., writing—review and editing, M.M., M.S., G.N., C.T., and A.V., visualization, M.M. and G.N. All authors have read and agreed to the published version of the manuscript.

## Data Availability

All data produced in the present work are contained in the manuscript

## Acknowledgements

Not applicable.

## Compliance with Ethical Standards/Disclosure of potential conflicts of interest

The authors have no conflicts of interest to declare that are relevant to the content of this article.

## Research involving Human Participants and/or Animals

Not applicable.

## Informed consent

Not applicable

