## Supplemental Table 1-5 for "Oxidative stress and antioxidant defenses in mild cognitive impairment: a systematic review and meta-analysis": ESF table.docx

**Running title: Oxidative stress in mild cognitive impairment**

Gallayaporn Nantachai, M.Sc.^a,b^, Asara Vasupanrajit, M.Sc.^a^, Chavit Tunvirachaisakul, M.D., Ph.D.^a,^ ^c^; Marco Solmi, M.D., Ph.D. ^d,e,f,g,h^, Michael Maes, M.D., Ph.D.^a,c,i,,j,^*

^a^ Department of Psychiatry, Faculty of Medicine, Chulalongkorn University, Bangkok, Thailand.

^b^ Somdet Phra Sungharaj Nyanasumvara Geriatric Hospital, Department of Medical Services, Ministry of Public health, Chon Buri Province, Thailand.

^c^ Cognitive Impairment and Dementia Research Unit, Department of Psychiatry, Faculty of

Medicine, Chulalongkorn University, Bangkok, Thailand.

^d^ Department of Psychiatry, University of Ottawa, Ontario, Canada.

e Department of Mental Health, The Ottawa Hospital, Ontario, Canada.

f Ottawa Hospital Research Institute (OHRI) Clinical Epidemiology Program University of Ottawa, Ottawa, Ontario.

^g^ Early Psychosis: Interventions and Clinical-detection (EPIC) Lab, Institute of Psychiatry, Psychology & Neuroscience, Department of Psychosis Studies, King's College London, London, United Kingdom.

^h^ Centre for Innovation in Mental Health-Developmental Lab, School of Psychology, University of Southampton, and NHS Trust, Southampton, United Kingdom.

^i^ IMPACT Strategic Research Center, Deakin University, Geelong, Australia.

^j^ Department of Psychiatry, Medical University of Plovdiv, Plovdiv, Bulgaria.

**Corresponding author:**

Prof. Dr. Michael Maes, M.D., Ph.D.

Department of Psychiatry,

Faculty of Medicine

Chulalongkorn University,

Bangkok

10330, Thailand

**ESF Table 1**. The nitro-oxidative / antioxidant (O&NS/ANTIOX) profiles examined in this study.

| ID Profile | Biomarkers | Function |
| --- | --- | --- |
| O&NS | 3NT + 7KC + oxLDL + AOPP + CP + Cu + dROM + F2A + Fe + GSSG + Hcy + HNE+ LOOH + MDA + NO + OHC + PGF2A + ROOH + ROS + TBARS | From ROS to O&NS |
| ANTIOX | Alb + BAP + CAT + CoQ10 + Cp + FRAP + Gpx + GR + GSH + HDL-c + MEL + Mg + Mn + PAO + RAP + Se + SOD + TAC + TAP + Tf + Thiols + vitB1 + vitB2 + vitB6 + vitB9 + vitB12 + vitA + vitC + vitD + vitE + Zn + Apig + Hesp + Kaemp + Lut + Lyc + Naring + Querc + Zeaxan | Total antioxidant capacity (endogenous + exogenous) |
| O&NS-toxicity | 3NT + 7KC + oxLDL + AOPP + CP + F2A + Fe + Hcy + HNE+ LOOH + MDA + NO + OHC + PGF2A + ROOH + TBARS + | Damage due to O&NS |
| Lipid-peroxidation | 7KC + oxLDL + F2A + HNE+ LOOH + MDA + OHC + PGF2A + ROOH + ROS + TBARS | Damage to lipids due to O&NS |
| Protein-oxidation | AOPP + CP | Damage to proteins due to O&NS |
| Homocysteine metabolism | Hcy - vitB9 | Increased homocysteine levels versus protective effects of vitB9 |
| Iron metabolism | Fe – Tf – Cp - FRAP | Iron-associated oxidative toxicity after considering the effects of Ferric Associated Antioxidant defenses |
| Non-vitamin ANTIOX | Alb + BAP + CAT + CoQ10 + Cp + FRAP + Gpx + GR + GSH + HDL-c + MEL + Mg + Mn + PAO + RAP + Se + TAC + TAP + Tf + Thiols + Zn | Non-vitamin-associated endogenous antioxidant capacity |
| Vitamin ANTIOX | vitB1 + vitB2 + vitB6 + vitB9 + vitB12 + vitA + vitC + vitD + vitE | Vitamin-associated antioxidant capacity |
| Glutathione system | GSH + Gpx + GR | Glutathione system associated antioxidant capacity |

**List of Abbreviations:**

3NT: 3-nitrotyrosine

8OHdG: 8-hydroxy-2'-deoxyguanosine

Alb: albumin

AOPP: advanced oxidation protein products

Apig: apigenin

BAP biological antioxidant potential

CAT: catalase

CoQ10: coenzyme Q10

Cp: ceruloplasmin

Cu: copper

dROM: diacron-reactive oxygen metabolite

F2A: isoprostane

Fe: iron

FRAP: ferric Reducing Antioxidant Power

Gpx: glutathione peroxidase

GR: glutathione reductase

GSH: glutathione, Reduced glutathione

GSHt: total GSH

GSSG: glutathione oxidized

Hcy: homocysteine

HDL-c: high-Density lipoprotein cholesterol

Hesp: hesperetin

HNE: hydroxynonenal

Kaemp kaempferol

LOOH: lipid hydroperoxide

Lut: luteineolin

Lyc: lycopene

MDA: malondiadehyde

MEL: melatonin

Mn: manganese

Mg: magnesium

Narin: naringenin

NO: nitric Oxide

OHC: hydroxycholesterol

oxLDL: anti-oxidized low-density-lipoproteins antibodies

PAO: total antioxidant power

PC: protein carbonyls

Quer: quercetin

RAP: residual antioxidant power

ROOH: hydroperoxides

ROS: reactive oxygen species

Se: selenium

SOD: superoxide dismutase

TAB: total antioxidant power

TAC: total antioxidant capacity

TBARS: thiobarbituric acid reactive substances

Tf: transferrin

Vit: vitamin

Zn: zinc

**ESF Table 2**. Specific search in each database

**PubMed/Medline**

((Oxidative stress)) AND (Antioxidants)) AND (blood biomarkers) OR (Lipids) OR (vitamin) AND (MCI)) **1564**

**PubMed/Medline**

((Oxidative stress OR Antioxidants OR Blood biomarkers) AND (Zinc) OR (Vitamin B) OR (Albumin) OR (SOD) OR (Lipid hydroperoxides) OR(CAT) OR (GSH) OR (Folic) OR (Coenzyme Q10) OR (Vitamin E) OR (Vitamin C) OR (Vitamin A) OR (Fe) OR (Hcy) OR (HDL) OR (Se) OR (Protein carbonyl) OR (Iron) AND (Mild cognitive impairment*) **83**

**Google scholar**

MCI AND Oxidative stress blood biomarkers OR Antioxidants AND Vitamins AND GSH AND Iron AND Hcy AND lipids **505**

**Scopus**

MCI AND blood* AND "oxidative stress biomarkers" AND antioxidants AND (LIMIT-TO (DOCTYPE, "ar")) **55**

**WEB OF SCIENCE**

(Mild cognitive impairment) (All Fields) and (Oxidative stress biomarkers) (Topic) **294**

**WEB OF SCIENCE**

(Mild Cognitive Impairment) AND ALL FIELDS: (oxidative stress biomarkers) AND ALL FIELDS: (Blood) AND ALL FIELDS: (Antioxidant) **28**

**ESF Table 3**. Oxidative stress confounders scale (OCS)

|  | **Methodological quality of the study** |
| --- | --- |
| 1 | Study sample ≥ 128 participants including patients and controls (1= Yes, 0= No) |
| 2 | Did the study control the results for potential confounders (e.g. age, BMI, gender, race)? (1= Yes, 0 = No) |
| 3 | Were participants with mild cognitive impairment (MCI) and controls age- and-gender-matched or statistically controlled for? (1= Yes, 0 = No) |
| 4 | Was the time of sample collection specified (e.g. morning vs. evening)? (1= Yes, 0 = No) |
| 5 | Were participants with MCI free of immunomodulatory drugs including anti-cytokines, glucocorticoids, immunoglobulins, and immunosuppressants, or been through a medication washout or intake statistically controlled for? (1= Yes, 0 = No) |
| 6 | Were participants with MCI free of acetylcholine esterase inhibitors (donepezil, rivastigmine, galantamine), memantine, nicergoline, ginkgo biloba extract) or were the data statistically controlled for the dug state? (1= Yes, 0 = No) |
| 7 | Reporting of either the manufacturer of the test or its parameters (detection limit and coefficient of variation) (1= Yes, 0 = No) |
| 8 | Where needed, reporting how data under detection limit was handled (1 = Yes, 0 = No) |
| 9 | Where needed, reporting % of the sample under detection limit (1=Yes, 0= No) |
| 10 | Reporting blood fraction (serum, plasma, culture supernatant or whole blood) (1= Yes, 0 = No) |
|  | **Total quality score (10 points)** |
|  | **Oxidative stress confounders red points**  *Red points should not be given if the item is statistically controlled for* |
| 1 | 3 red points for comorbid illnesses such as autoimmune disorders & other immune disorders including RA, psoriasis, IBD, COPD, MS |
| 2 | 3 red points for use of recreational drugs such as methamphetamine or opioids (not applicable if all axis-1 psychiatric disorders are excluded) |
| 3 | 2 red points for comorbidity with MDD / BD (not applicable if all axis-1 psychiatric disorders are excluded) |
| 4 | 2 red points when groups were not matched or controlled for age |
| 5 | 2 red points when groups are not matched or controlled for sex |
| 6 | 2 red points for medication use as for example immunomodulators |
| 9 | 1.5 red points for antidepressants |
| 10 | 1 red point for other neuro-psychiatric comorbidities, as for example schizophrenia, autism, GAD, PTSD |
| 11 | 1 red point for more common systemic immune disorders including diabetes type1/2, essential hypertension, metabolic syndrome |
| 12 | 1 red point for not fasting (8 hours before blood extraction) |
| 13 | 1 red point for use of omega-3 and antioxidant supplements |
| 14 | 1 red point when groups were not controlled for BMI |
| 15 | 1 red point when groups were not controlled for physical activity or sedentary life |
| 16 | 1 red point when groups were not adjusted for smoking  1 red point when groups were not adjusted for alcohol use |
| 18 | 0.5 red points when groups were not controlled for ethnicity (in countries such as US, Brazil |
| 19 | 0.5 red points when there was no adjustment for seasonality |
| 20 | 0.5 red points when groups were not matched for diurnal variation (8-10 a.m. versus all other time points) |
|  | **Total red score (25 points)** |

Adapted from Andrés-Rodríguez et al., 2019; Vasupanrajit et al., 2121;2021

*This threshold of study samples is established as it is the minimum needed for a statistical power of 0.8.

Oxidative stress cofounders red points should be given when the item is not reported (or statistically controlled for).

**ESF Table 4**. Studies excluded from the Meta-Analysis but included in the systematic review.

| Authors, year | Reason why excluded from  the Meta-Analysis | Results |
| --- | --- | --- |
| Rozzini et al., 2018 | MCI due to Alzheimer’s disease (ADMCI). | Free and total serum copper levels were significant different (p˂0.02) between ADMCI and healthy controls,  There was a significant difference in free copper (p=0.001) in ADMCI as compared with healthy controls. |
| Vinothkuma et al., 2017 | MCI comorbid with chronic kidney disease (CKD) | Plasma SOD, CAT, Gpx, and reduced GSH were significantly decreased in CKD with cognitive dysfunction as compared with CKD without cognitive dysfunction, and lipid peroxidation was significantly increased in CKD with cognitive dysfunction *versus* CKD without cognitive dysfunction. |
| Zheng et al., 2019 | MCI comorbid with type 2 diabetes (T2DM) | There was a significant difference in serum folate (p<0.001) in T2DM-MCI *versus* controls, but no significant difference in HDL-c |

**ESF Table 5.** Description of all studies included in the Meta-Analysis

| Authors,  years | Setting | Criteria* | N | | Assessed Biomarkers | Specimen | Exclude comorbid illness | Med wash out | Quality score | Red point score | Results |
| --- | --- | --- | --- | --- | --- | --- | --- | --- | --- | --- | --- |
|  |  |  | MCI | HC |  |  |  |  |  |  |  |
| AI-Rawaf et al., 2021 | Saudi Arabia | Yes | 70 | 80 | MDA, CAT, SOD, NO | Plasma Hemoglobin | Partly | Partly | 5.25 | 14.50 | MCI > HC***: MDA, NO  MCI < HC***: CAT, SOD |
| Arce-Varas et al., 2017 | Spain | Yes | 43 | 44 | SOD, Gpx, CAT | Plasma | No | Partly | 4.75 | 19.25 | MCI < HC*: SOD  MCI > HC***: GPx |
| Ayromlou et al., 2018 | Iran | Yes | 45 | 45 | VitC, VitA, Lyc | Serum | Yes | Yes | 4.5 | 14 | MCI < HC *: VitA, Lyc |
| Balmus et al., 2017 | Romania | Yes | 15 | 15 | SOD, Gpx, MDA, Mg, Mn, Fe | Serum | Partly | No | 4.25 | 18 | MCI < HC ***: SOD, Gpx, MDA  MCI < HC *: Mg  MCI > HC*: Mn |
| Bermejo et al., 2008 | Spain | Yes | 34 | 28 | PC, GSH, GSSG, GSH/GSSG, Gpx | Plasma, Erythrocytes | Partly | Partly | 4 | 19 | MCI > HC*: PC MCI < HC*: GSSG |
| Boccardi et al., 2021 | Italy | Yes | 95 | 65 | HDL-c, VitB12, VitB9 | Serum | Yes | Yes | 5 | 16.5 | MCI > HC: HDL  MCI < HC: VitB12, B9 but NS between group |
| Cervellati et al., 2014 (a) | Italy | Yes | 103 | 48 | AOPP, Hcy, LOOH, Thiol, TAP, RAP | Serum | Partly | Partly | 6 | 16 | MCI > HC*: LOOH  MCI < HC*: RAP |
| Cervellati et al., 2013 (b) | Italy | Yes | 134 | 99 | HCL-c, Alb, LOOH, AOPP, TAP, RAP, Thiol | Serum | Yes | Partly | 6 | 15.5 | MCI < HC*: RAP |
| Cervellati et al., 2014 (c) | Italy | Yes | 82 | 118 | LOOH, RAP | Serum | Yes | Partly | 6 | 16 | MCI > HC*: LOOH |
| Chico et al., 2013 | Italy | Yes | 52 | 63 | AOPP, FRAP | Plasma | No | No | 3 | 18.5 | MCI > HC***: AOPP  MCI < HC***: FRAP |
|  |  |  | 20 | 21 | SOD |  |  |  |  |  |  |
| Chmatalova et al., 2017 | Creech Republic | Yes | 17 | 12 | Se | Plasma | Yes | No | 3 | 17.5 | MCI < HC**: Se |
| Dong et al.,  2009 | USA | Yes | 19 | 16 | Zn | Serum | No | No | 3.5 | 21 | MCI < HC: Zn |
| Du et al.,  2019 | China | Yes | 113 | 832 | Alb, dROM, BAP, BAP/ dROM ratio | Serum | No | Partly | 5.5 | 13.5 | MCI > HC***: dROM  MCI < HC***: BAP, BAP/dROM ratio |
| Gironi et al., 2011(a) | Italy | Yes | 20 | 66 | MDA, GStot GSH, PAO | Serum | partly | Partly | 3.5 | 19 | MCI < HC*: GSH  MCI > HC*: PAO |
| Gironi et al., 2015 (b) | Italy | Yes | 26 | 28 | PAO, oxLDL, ROS, CoQ10, MDA, GSHt, GSH, GSSH | Plasma, serum | No | No | 4.25 | 17.75 | MCI ˂ HC*: GSH  MCI > HC***: GSSH |
| Guidi et al.,  2006 | Italy | Yes | 36 | 44 | Hcy, ROS, TAC | Plasma | No | no | 2.5 | 20 | NS between group |
| Gunduztepe et al., 2020 | Turkey | Yes | 19 | 30 | Alb, Cp, thiols | Serum | Yes | Partly | 3.5 | 21 | MCI >HC*: Cp |
| He et al., 2016 | China | Yes | 112 | 115 | HDL-c | Plasma | Yes | No | 3 | 16 | MCI < HC*: HDL |
| Irizarry et al., 2007 | USA | Yes | 47 | 48 | F2A | Plasma | No | No | 4 | 16 | NS between group |
| Iuliano et al., 2010 | Italy | Yes | 24 | 24 | VitE, OHC | Plasma | Partly | Partly | 3.75 | 11.25 | NS between group |
|  |  |  | 29 | 24 | VitE, OHC |  |  |  |  |  |  |
| Kim et al., 2013 | South Korea | Yes | 100 | 121 | VitB9, VitB12, Hcy | Plasma | Partly | No | 4.5 | 16 | MCI > HC***: Hcy |
| Lanyau-Dominguez  et al., 2020 | Cuba | Yes | 126 | 214 | Hcy, VitB1 | Plasma | Yes | No | 3.5 | 16.5 | MCI > HC***: Hcy |
|  |  |  | 120 | 215 | VitB2 | Plasma | Yes | No | 3.5 | 16.5 | MCI > HC*: VitB2 |
|  |  |  | 126 | 228 | VitB9, VitB12 | Serum | Yes | No | 3.5 | 16.5 | NS between group |
|  |  |  | 122 | 226 | VitC | Plasma | Yes | No | 3.5 | 16.5 | MCI < HC**:  Vit C |
|  |  |  | 111 | 197 | VitA | Plasma | Yes | No | 3.5 | 16.5 | MCI < HC**: VitA |
| Lee et al., 2013 | Malaysia | Yes | 67 | 134 | HDL-c | Serum | Partly | No | 5.25 | 8.25 | NS between group |
| Lopez et al., 2013 | Spain | Yes | 18 | 33 | Cu, MDA, SOD, Cp | Plasma, Serum | Partly | No | 3.5 | 20 | MCI > HC***: MDA  MCI > HC*: Cu |
| Lui et al., 2016 | China | Yes | 70 | 140 | HDL-c, OHC | Plasma, Serum | Yes | No | 4 | 10.5 | MCI > HC**: OHC |
| Ma et al., 2017 | China | Yes | 112 | 115 | Hcy, VitB9, VitB12 | Plasma, Serum | Partly | No | 5.5 | 13 | MCI˂HC*: VitB9, VitB12  MCI > HC**: Hcy |
| Martin-Aragon et al., 2009 | Spain | Yes | 34 | 28 | Gpx-Se, GR, MDA | Plasma | Partly | Partly | 4 | 19 | NS between group |
| Martinez de Toda et al.,  2019 | Spain | Yes | 20 | 30 | SOD, CAT, Gpx, GR, GSH, GSSH, GSH/GSSG, TBARS | Erythrocytes | partly | no | 3.5 | 19 | MCI < HC***: GPx, GR  MCI ˃ HC***: CAT, TBARS  MCI > HC**: GSH/GSSH |
| McFarlane et al., 2020 | Poland | Yes | 85 | 71 | OHC, HDL-c | Plasma, Serum | Yes | No | 4 | 13.5 | NS between group |
| Mota et al., 2015 | Portugal | Yes | 24 | 20 | Basal and induced ROS, | Serum | Partly | No | 2.5 | 16.5 | MCI > HC*: Basal ROS MCI˃HC*: Induced ROS |
| Mueller et al., 2012 | USA | Yes | 7 | 19 | Cu, Fe | Serum | No | No | 2.5 | 19 | At 1-year prior progression to AD  St-MCI, pr-MCI> HC*: Cu, Fe |
|  |  |  | 8 | 19 | Cu, Fe |  |  |  |  |  |  |
| Mufson et al., 2010 | USA | Yes | 34 | 167 | F2A | Plasma | No | No | 4 | 20 | NS between group |
| Negahdar et al., 2015 | Iran | Yes | 57 | 120 | FRAP, TBARS, Zn, Cu, Mn, Hcy, | Serum | No | Partly | 5.5 | 15 | MCI I < HC***: FRAP  MCI I> HC***: TBARS  MCI I> HC**: Hcy |
|  |  |  | 63 | 120 | FRAP, TBARS, Zn, Cu, Mn, Hcy, | Serum | No | Partly | 5.5 | 15 | MCI II< HC***: FRAP  MCI II > HC***: TBARS  MCI II > HC**: Hcy |
| Padurariu et al., 2010 | Romania | Yes | 15 | 15 | SOD, Gpx, MDA | Serum | Partly | No | 4.25 | 18.25 | MCI < HC***: SOD  MCI < HC****: GPx  MCI > HC***: MDA |
| Perrotte 2019 | Canada | Yes | 24 | 24 | Alb, VitD, HDL-c | Plasma | Yes | Partly | 4 | 12.5 | NS between group |
| Praticó et al., 2002 | USA | Yes | 33 | 40 | F2A | Plasma | Partly | No | 2.75 | 16 | MCI > HC***: F2A |
| Rembach et al., 2013 | Australia | Yes | 96 | 716 | Cu, Cp | Serum | No | No | 4.5 | 21.5 | At 8-month MCI > HC*: Cp |
| Rinadi et al., 2003 | Germany | Yes | 25 | 56 | VitC, VitE, VitA, Lut, Zeaxan, Lyc, SOD, Gpx | Plasma, RBC | Yes | No | 3.5 | 15 | MCI < HC****: VitC, VitE, SOD, Gpx  MCI < HC***: Vit A  MCI < HC**: LUT, Zeaxan, VitA |
| Rita Cardoso et al., 2014 | Brazil | Yes | 52 | 29 | Se, Lipids | Plasma, Erythrocytes | No | No | 4 | 16.75 | MCI < HC **: Se |
| Sirin et al.,  2015 | Turkey | Yes | 21 | 22 | F2A, MEL | Plasma | No | Partly | 5.25 | 10.5 | NS between group |
| Squitti et al., 2011 | Italy | Yes | 83 | 100 | Cu, free Cu, Cp, TRAP, Tf, Fe | Serum | No | Partly | 6 | 10.75 | MCI > HC**: Cu, Free Cu |
| Sultana et al., 2013 | Italy | Yes | 5 | 5 | PC, HNE, 3NT | Mitochondria | No | No | 3 | 18 | MCI > HC*: PC, HNE, 3NT |
| Torres et al., 2011 | Brazil | Yes | 33 | 26 | MDA, CAT, Gpx,GR, GR/GPx | Plasma, Erythrocytes | Yes | Partly | 4.75 | 18 | MCI < HC***: GR/GPx  MCI > HC*: MDA  MCI > HC**: GR |
| Ulstein et al., 2017 | Norway | Yes | 25 | 63 | Hcy, VitB1, VitB6, VitB9, VitB12, VitC, VitA, VitE, VitD, F2A | Serum | Yes | No | 3.75 | 16.5 | NS between group |
| Yuan et al., 2016 | China | Yes | 138 | 138 | HDL-c, TAC, MDA, GSH, SOD, CAT, Gpx, GSHt, GR, VitA, VitE, Lut, Quer, Narin, Apig, Kaemp, Hesp | Plasma,  Erythrocytes | Partly | No | 5.5 | 14 | MCI < HC***: HCL  MCI < HC***: VitE  MCI < HC**: TAC  MCI > HC*: MDA |
| Zhou et al.,  2019 | China | Yes | 118 | 118 | VitB9, VitB12, Hcy | Serum | No | No | 4 | 15.5 | MCI < HC*: VitB12  MCI > HC*: Hcy |

Significant differences between groups at *p˂0.05, ** p˂0.01, *** p˂0.001, **** p˂0.0001, no significant differences (NS)

Total quality score is maximal 10 points; Total red point score is maximal 25 points

For list of abbreviations and O&NS/ANTIOX profile description, see ESF Table 1

Criteria*: studies reporting on MCI/aMCI as diagnosed using Petersen criteria by clinicians or using neuropsychology test results

All the studies reviewed here used healthy volunteers as control group
